# Epidemiological study to detect active SARS-CoV-2 infections and seropositive persons in a selected cohort of employees in the Frankfurt am Main metropolitan area

**DOI:** 10.1101/2020.05.20.20107730

**Authors:** Verena Krähling, Martin Kern, Sandro Halwe, Helena Müller, Cornelius Rohde, Massimiliano Savini, Michael Schmidt, Jochen Wilhelm, Stephan Becker, Sandra Ciesek, René Gottschalk

## Abstract

So far, 170,000 *Severe acute respiratory syndrome-related coronavirus 2* (SARS-CoV-2) infections have been confirmed in Germany, of which more than 5,000 have been detected in the Frankfurt am Main metropolitan region. When examining 1,000 nasopharyngeal swabs and serum samples from healthy volunteers from this region, one RT-PCR-positive and five antibody-positive persons were identified. The five positive serum samples were confirmed to be specific. Four of the five positive sera cross-neutralized SARS-CoV.

## Main Text

More than 4 million people have been infected with SARS-CoV-2 globally with nearly 280,000 fatalities. It is important for the containment of the SARS-CoV-2 pandemic to be aware of the total number of infected people, which is probably underestimated due to the underreporting of mild and asymptomatic cases. In preliminary work we examined symptom-free travelers for SARS-CoV-2-specific RNA to detect unnoticed COVID-19 cases. Among 122 passengers, who returned from Wuhan in February 2020, we identified two infected asymptomatic individuals (data not shown). This result suggests that the screening of asymptomatic individuals could be helpful in estimating the actual number of acute infections in the population. Several reports indicated that a combination of Realtime PCR (RT-PCR) and serological assays can be superior to identifying SARS-CoV-2 compared to RT-PCR alone [1–3].

The project aims to determine the current SARS-CoV-2 status of a selected cohort of 1,000 employees in the Frankfurt metropolitan area to contribute to an accurate knowledge of the status of the SARS-CoV-2 pandemic.

### Description of the study

The study was announced to the employees of Infraserv Hochst, a large industrial site operator in Frankfurt am Main at the beginning of April 2020 by the company’s own Occupational Health Center. All employees were allowed to participate until a cohort of 1,000 subjects was reached. There were no exclusion criteria. Nasopharyngeal swab and blood samples (serum) were taken from participants between April 6th and April 14th. In case of proven SARS-CoV-2 infection, the participant and the health authorities were informed immediately. This study was carried out in accordance with the Declaration of Helsinki and the guidelines of the International Conference for Harmonization for Good Clinical Practice. The study was approved by the local Ethics Committee at the University Hospital Frankfurt. Informed consent was waived due to the retrospective character of the study. All participants have given their written consent.

21.5% of the participants were women and 78.5% were men, all aged between 18 and 65 years (Table 1). The distribution of the participants’ places of residence is shown in Figure 1. The participants showed no symptoms prior to the tests. Two participants stated that they had a SARS-CoV-2 infection at the beginning of March 2020. The number of reported SARS-CoV-2 infections in the districts where the study participants live was 5,177. The total number of inhabitants is approximately 5 million.

**Table 1.**
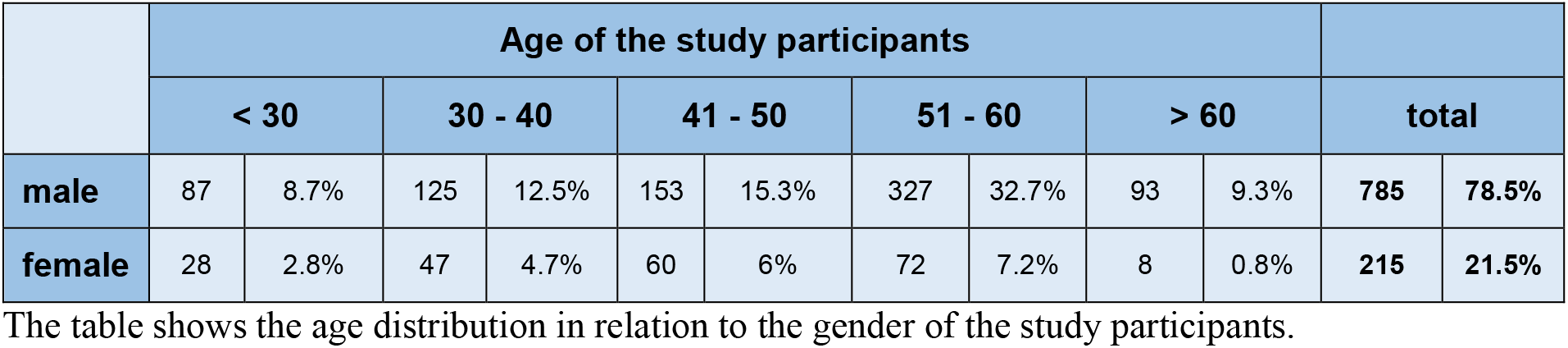
The table shows the age distribution in relation to the gender of the study participants.

**Figure 1.**
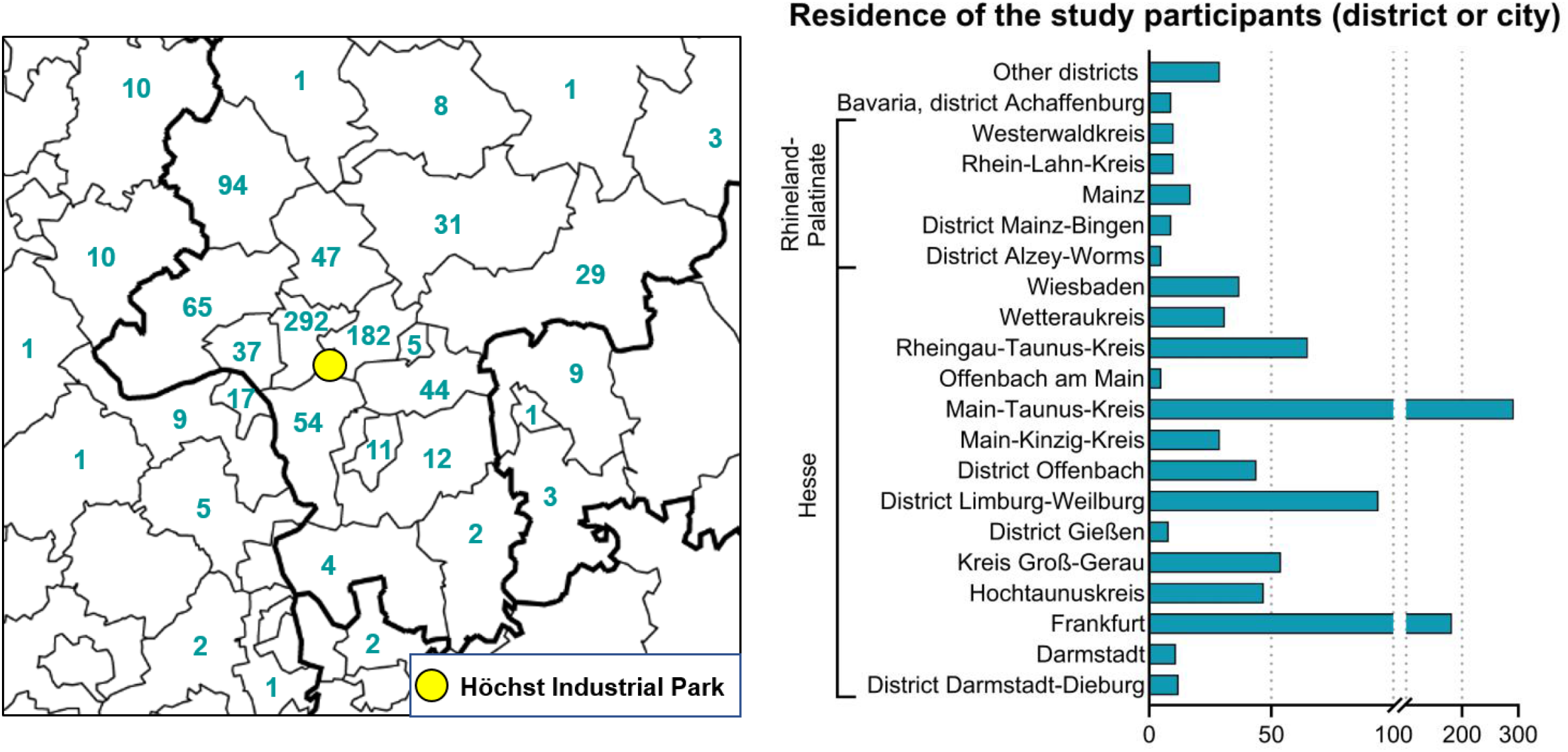
Distribution of the participants’ places of residence

Most participants live in the larger Frankfurt am Main metropolitan area

### Screening for active SARS-CoV-2 infections by RT-PCR

RT-PCR was performed on the Roche Cobas® 8800 instrument. The sample input volume was 400 μl. Amplification was done in a multiplex CE certified assay in the ORF region as well as in the E-gene. All samples were tested in accordance to the instruction for use from the manufacturer (Roche Diagnostics, Mannheim, Germany). Among 1,000 individuals screened, we found one person positive for SARS-CoV-2. This person had the nasopharyngeal swab taken on April 9th. The finding was reported to the volunteer on April 12th. No symptoms were recorded at this time. On April 14th a sudden loss of taste in the early morning was described that persisted for 24 hours. On April 15th to 16th a mild headache was reported, from April 17th on there were no further symptoms recorded.

### Discovery of individuals who seroconverted unnoticed

Serological assays have been published that are based on the recombinant SARS-CoV-2 proteins N, S or truncated versions of the S protein (S1) for the detection of antibodies against SARS-CoV-2 [1–7]. Although the results vary, the studies agree that the S1-based tests are most specific for SARS-CoV-2 and have the least cross-reactions with other coronaviruses [4, 5]. For the development of an *in-house* ELISA for the detection of anti-SARS-CoV-2 antibodies we used 100 ng recombinant SARS-CoV-2 S1 per well (Sino Biological, 40591-V08H) in combination with an HRP-coupled anti-human IgG secondary antibody. Serum samples were analysed once at a dilution of 1:100. Samples that exceeded an OD of 0.2 were re-analysed in duplicates. Those that exceeded an OD of 0.3 in the re-analysis were classified as positive. The final cut-off value of the ELISA was determined by analysing the OD values of 120 negative sera from 2014 with no history of SARS-CoV infection. The cut-off value was calculated as the average of the OD values plus 4 standard deviations. Classifying the negative samples according to the procedure described above resulted in one false-positive, indicating a specificity of 99.2% (95% confidence interval (CI): 95.4% - 99.98%). The analysis of successive sera from fifteen SARS-CoV-2-PCR-positive individuals showed seroconversion in all cases (data not shown). The sample size is not large enough to make any precise statement about the sensitivity (95% CI: 87,2% to 100%).

Using the newly developed SARS-CoV-2 ELISA, 1,000 sera of volunteers were screened for the presence of SARS-CoV-2 S1 binding antibodies (Figure 2). We identified 29 reactive sera (Figure 3A, bars), which were further analysed for their neutralizing capacity against SARS-CoV-2. Briefly, sera were serially diluted starting with a dilution of 1:8. Then, 100 TCID_50_ units of SARS-CoV-2 (European Virus Archive Global # 026V-03883) were added. Following incubation at 37 °C for 1 h, Vero C1008 cells (ATCC CRL-1586) were added per well. Cytopathic effect (CPE) was evaluated at day 4 post infection. Neutralization titers of four replicates were calculated as geometric means (reciprocal value).

**Figure 2.**
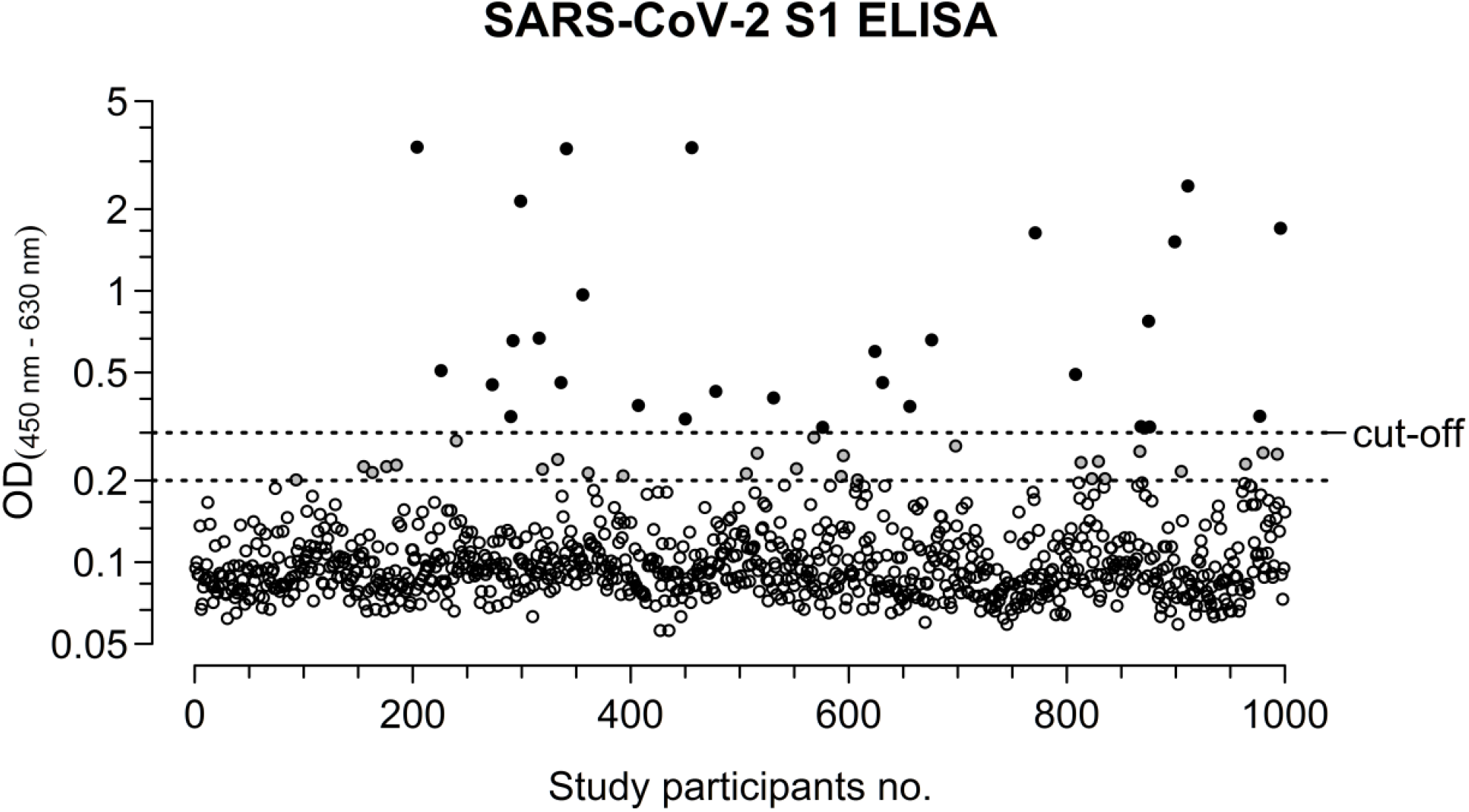
SARS-CoV-2 spike (S1) protein ELISA

**Figure 3.**
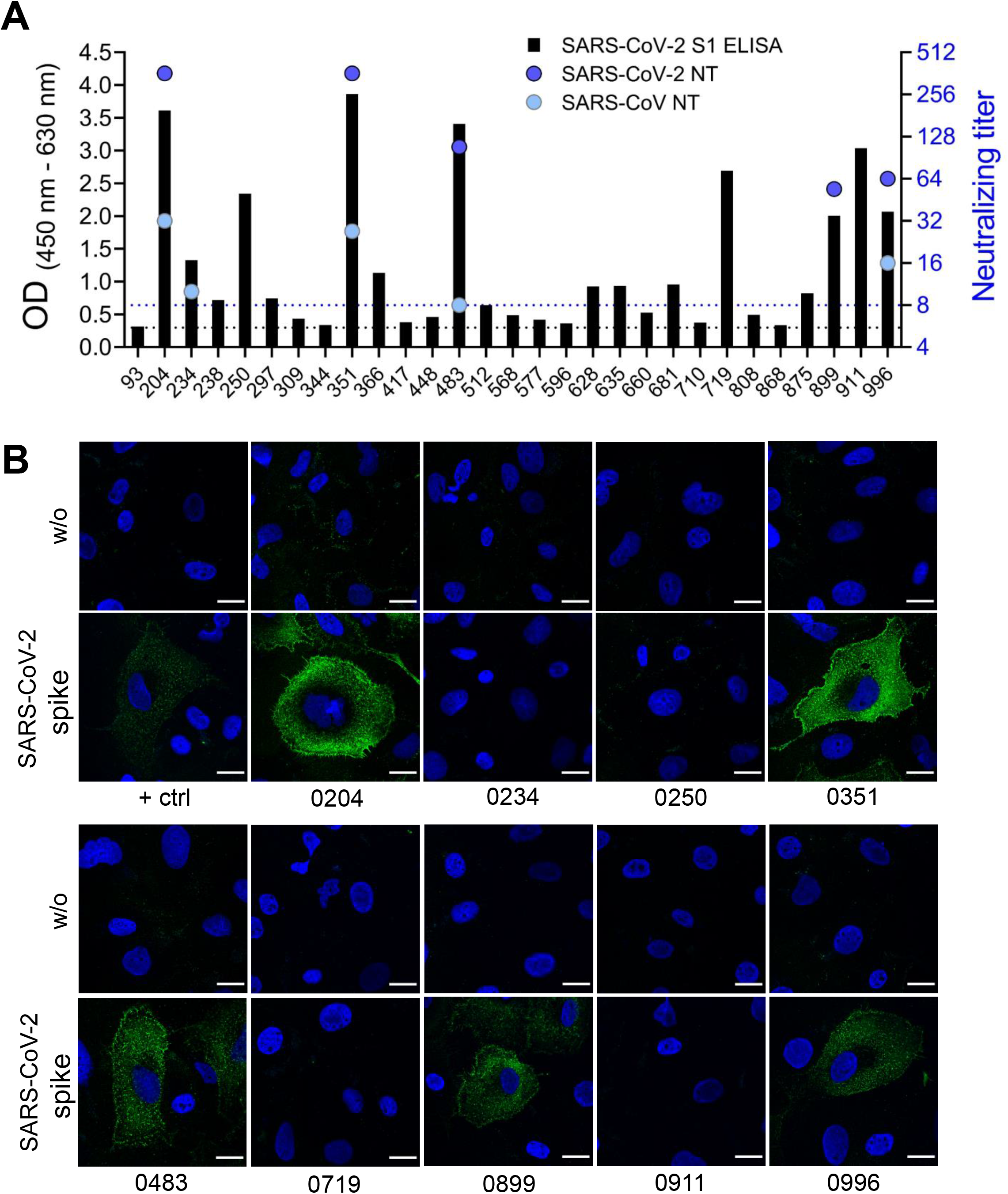
Reactivity of serum antibodies on SARS-CoV-2 or SARS-CoV antigen

The figure shows the results of the screening of 1,000 serum samples in a SARS-CoV-2 spike (S1) ELISA.

Among the 29 reactive sera, SARS-CoV-2 neutralising antibodies were detected in five sera (Figure 3A). These sera were also reactive to the spike protein at the surface of S1 expressing cells by native immunofluorescence analysis (nIFA as described in Krähling *et al*., 2016 [10], with the exception that a pCAGGS-SARS-CoV-2 spike expression plasmid was used) (Figure 3B). The remaining 24 sera were not reactive in either assay. We conclude that five out of 1,000 individuals tested developed SARS-CoV-2-specific antibodies. The remaining 24 individuals are likely to have cross-reactive antibodies from other human coronavirus infections. The five sera that were considered truly positive were reactive in the nIFA and in the neutralization test (NT), which indicated that it is possible to confirm the ELISA results with nIFA. This is an important result for laboratories that do not have access to a BSL3 facility.

**A** Characterization of 29 of the 1,000 serum samples, which were tested positive in the SARS-CoV-2 spike (S1) ELISA (black bars). The neutralization of SARS-CoV-2 by the respective serum is shown by the dark blue circles, the neutralization of SARS-CoV is shown by the light blue circles. **B** Representative images of nIFA. Microscopic analyses were performed at a magnification of 63x with a confocal laser scanning microscope (Leica SP5). All images were acquired with a laser intensity of 10 % (488 nm excitation). Scale bar: 20 μM.

Two of the five seropositive volunteers stated that they underwent a laboratory-confirmed SARS-CoV-2 infection (PCR-based). The other three subjects had not been aware of the infection. The first subject of this group had no symptoms in the past months and no known contact with a COVID-19 patient. The second subject had common cold symptoms 7 weeks before the start of the study, which was assumed to be caused by pollen allergy. This individual went to work regularly in overall healthy condition. The third subject had contact with a colleague who had subsequently tested positive for SARS-CoV-2. As a precaution he stayed at home for two weeks. At no time did he experienced any symptoms.

The 29 serum samples reactive in the SARS-CoV-2 ELISA were further analyzed using a SARS-CoV (2003) neutralisation test (FFM-1 isolate, GenBank accession number AY310120). The assay was performed as described for SARS-CoV-2 with the exception that the CPE was evaluated at day three post infection. Four out of five sera that bound the SARS-CoV-2 S1 protein and neutralised the SARS-CoV-2 virus also neutralised SARS-CoV. In contrast to published results [11], we do see a weak cross-neutralization of SARS-CoV by SARS-CoV-2-specific antisera. This can be due to the use of different test systems. Ou and colleagues used pseudotyped V SV for their neutralization tests while we used wild-type SARS-CoV-2 and SARS-CoV.

## Conclusion

Most of the officially recorded COVID-19 cases showed symptoms or had relevant contact with positively tested index patients. In contrast, among the 998 tested employees with unknown SARS-CoV - 2 status, we found four who did not know they were acutely or previously infected with SARS-CoV-2 (95% credibility interval (CrI): [0.163%; 1.022%]; Bayesian analysis using a flat prior [12]). This result indicated the rate of unreported to reported SARS-CoV-2 cases in this study group was 2, a value which was unexpectedly low when compared with other respiratory diseases such as influenza (underreporting factor 79, USA, flu pandemic 2009, [13]). One limitation of our study is that the sample was not random because only volunteers were included, so the sample may not be representative. A SARS-CoV-2 surveillance study, performed in Heinsberg, North Rhine Westphalia, Germany, reported 15.5% SARS-CoV-2-infected individuals in a group of 919 [14]. The site of this study was a hot spot of SARS-CoV-2 transmission. The different SARS-CoV-2 prevalences (0.6% Frankfurt vs 15.5% Heinsberg) reflect the inhomogeneous pattern of the SARS-CoV-2 pandemic in Germany. To provide a reliable estimation of the average rate of SARS-CoV-2 infections in the general population in Hesse or Germany it is therefore mandatory to perform studies across the entire country and to combine these data. It will be also necessary to perform follow-up serological studies to determine the further spread of SARS-CoV-2.

## Data Availability

The data that support the findings of this study are available from the corresponding author, upon reasonable request.

